# Classical machine learning versus transformers for predicting daily psychological distress from passive smartphone sensing and self-report: a prospective cohort study

**DOI:** 10.64898/2026.11.04.26367218

**Authors:** Ike Griss Salas, Wai-yin Lam, Robyn E. Kilshaw, Brian R. W. Baucom, Nina de Lacy

## Abstract

**Objective:** We test whether data collected from daily passive smartphone sensing and self-reports can predict daily significant psychological distress with meaningful potential clinical utility, comparing across best-in-class machine learning algorithms.

**Methods:** In this prospective cohort study, 169 community-dwelling adults completed baseline surveys followed by 7 days of passive smartphone sensing (accelerometer, GPS, power state) and end-of-day Brief Symptom Inventory-18 (BSI-18) assessments. Cases were defined as days (total *n* = 1,062) where participants’ psychological distress measured with the sex-specific BSI-18 Global Severity Index T-score was ≥63 (prevalence 11.2%). Four classical models (artificial neural network, elastic net, support vector machine [SVM], extreme gradient boosting [XGB]) and four transformers (sensor-only or multimodal; standard or fusion architecture) were tuned, Platt-calibrated and evaluated on held-out participants (210 records, 25 cases) with participant-level bootstrap 95% confidence intervals (CIs).

**Results:** The SVM (area under the receiver operating characteristic curve [AUROC] 0.84; 95% CI 0.71–0.93), the multimodal fusion transformer (0.84; 0.69–0.95) and XGB (0.82; 0.63–0.94) discriminated best; sensor-only transformers performed at chance (0.49–0.50). At the default threshold of 0.50, elastic net logistic regression was the best classifier (sensitivity 0.56, positive predictive value 0.64). At 0.12 (near the outcome prevalence), the SVM detected all cases (negative predictive value 1.00), but positive predictive values were low for all models (0.13–0.26). Decision-curve analysis favored most models over treat-all and treat-none lines at thresholds between 0.05 and 0.25. Baseline survey items, mainly on functional impairment, had the largest feature importance.

**Conclusions:** Compact classical models performed as well as transformers, which were considerably more computationally intensive. Decision-curve analysis supports further clinical exploration of these tools for triaging daily psychological distress, though our results suggest the relative value of psychosocial assessments and digital data should be carefully analyzed. External validation is needed before clinical use.

## Introduction

Psychological distress is a state of emotional suffering that combines symptoms of depression, anxiety, and somatic complaints. It is common and often precedes or accompanies mental disorders, which account for about 15% of years lived with disability worldwide.^1^ Many affected people are not identified or treated. A median of 56% of people with depression receive no treatment,^2^ and in primary care only 47% of patients with depression are correctly identified.^3^ Brief self-report scales, such as the Kessler Psychological Distress Scale^4^ and the Brief Symptom Inventory-18 (BSI-18),^5,6^ are reliable screening instruments but require active and repeated completion, limiting continuous monitoring outside clinical settings. There is growing interest in using machine learning and artificial intelligence to advance the assessment of psychological states from both active (self-report) and passive (sensor) data.^7^

Digital phenotyping uses data from personal digital devices to measure behavior continuously in daily life.^8^ Smartphones record passive signals such as mobility (global positioning system [GPS]), physical activity (accelerometer), and phone use (screen and power state) with minimal effort from the user,^9^ complementing repeated self-reports with near-continuous behavioral data. Correlational studies have linked these signals to symptoms of depression.^10,11^ However, findings have been mixed: some signals showed contradictory associations across studies,^11^ and correlations in a larger sample were weaker than in earlier, smaller studies.^12^ Wearable sensor data also contributed little to predicting depression and suicidal ideation compared with daily self-report,^13,14^ and smartphone data alone predicted symptoms poorly until daily survey responses were added.^12^ It is therefore unclear whether passive sensor data are informative on their own or mainly in combination with self-report, and whether they have screening utility.

Systematic reviews attribute these gaps partially to small samples, short follow-up periods, and inconsistent measures,^11,15^ with models from small, homogeneous samples often failing to generalize.^16,17^ Analytic choices may also contribute. Most studies use correlation, regression or classical machine learning, which require fixed inputs, so time series are usually reduced to daily or weekly averages; models using the raw time series remain uncommon.^15,18^ Such averages can obscure when and how behavior changed over time.^18^ In addition, sensor data are often incomplete, and missingness itself correlates with participant engagement and symptom severity,^19,20^ so imputation may disguise behavioral changes.^18^ Many studies also split observations rather than participants for validation, which can greatly overestimate accuracy.^21^

Deep sequence models can learn directly from the sensor time series. Transformers^22^ use attention mechanisms to model temporal dependencies and have been adapted for time-series data^23^ and multimodal fusion.^24^ However, transformers typically need large training datasets.^23^ In clinical prediction, machine learning has shown no benefit over logistic regression at low risk of bias,^25^ and tree-based ensembles often outperform deep networks on medium-sized tabular datasets.^26^ Whether the extra complexity of transformers improves prediction at sample sizes typical in mental health research remains unclear. Many studies also report only discrimination,^25^ although reporting guidance recommends also evaluating calibration and clinical utility.^27–29^ Knowing which inputs drive a prediction can be as important as accuracy.^30^

We used data from a prospective cohort of community-dwelling adults who completed baseline surveys, 7 days of passive smartphone sensing and end-of-day BSI-18 assessments.^31^ Our objective was to determine whether new transformer models offer advantages over established classical machine-learning models for predicting daily clinically significant psychological distress from combined active (self-report) and passive (smartphone sensor) data, in terms of overall predictive performance and feature interpretability. To this end, we evaluated four classical models and four transformer configurations for discrimination, calibration, threshold-dependent classification and clinical utility, compared sensor-only with combined sensor and survey input to test whether passive sensor data were informative on their own, and examined which survey and sensor features drove the predictions.

## Methods

### Overview

This study compares classical machine learning models with transformer architectures for predicting daily psychological distress from combined passive smartphone sensor data and baseline self-report surveys. Transformers were evaluated because they can model temporal dependencies and natively fuse heterogeneous data modalities, capabilities that classical models applied to summary statistics do not offer. Because the binary distress outcome exhibited substantial within-person stability, all data partitioning and bootstrap resampling were performed at the participant level to prevent information leakage. The sections below describe the study sample and data sources, the predictive modeling pipeline, and the evaluation framework. All analyses were implemented in Python, mainly using scikit-learn, XGBoost, and PyTorch.

### Study procedures and participants

The sample consisted of 400 Utah-based participants recruited using a stratified random sampling design with two crossed factors—difficulties in emotion regulation and perceived life stress—to balance participants at high and low risk of episodic mental health conditions.^31^ Eligibility required age 18 years or older, Utah residency, smartphone ownership, English fluency, and health care through University of Utah Health or Intermountain Health; exclusions included suicidal ideation (past 3 months) or any suicide attempt, active mania or psychosis, or certain violence histories.^31^ After completing baseline questionnaires on psychiatric, medical, and psychosocial history, participants entered a 7-day in situ phase, providing continuous passive smartphone sensor data and daily self-report measures.^31^ Data were collected between June 2022 and June 2024. Of the 400, 253 provided sensor data; the analysis was restricted to 189 iPhone users by excluding 64 Android users for device consistency.

### Study data

Passive smartphone sensor data were collected using the Beiwe Research Platform.^32^ Accelerometer data recorded 3-axis acceleration at 10 Hz during 60-second on-cycles alternating with 60-second off-cycles. GPS data recorded latitude, longitude, and altitude, nominally sampled at 1 Hz on a 60-second on / 600-second off cycle.^19^ Power state data were event-based, logging screen lock/unlock transitions, charging state, and battery level.

Baseline questionnaire data were collected 4–209 days before the sensor monitoring period. The measures are described in the study protocol^31^ and in Supplementary Table 1. Features came from validated measures of emotion regulation (Difficulties in Emotion Regulation Scale [DERS]),^33^ perceived stress (Perceived Stress Scale-4 [PSS-4]),^34^ depression, anxiety, and stress symptoms (Depression Anxiety Stress Scales [DASS-21]),^35^ substance use risk (Tobacco, Alcohol, Prescription Medication, and Other Substance Use Tool [TAPS]),^36^ and psychosocial functioning (Life Functioning Questionnaire [LFQ]),^37^ as well as questionnaires assessing demographics, clinical history, and medical conditions.^31^ Measures were scored following published guidelines; all item-level responses and relevant subscale and total scores were included as features (260 in total).

The sample size was determined by participants meeting inclusion criteria rather than an *a priori* power calculation. After applying the outcome-based inclusion criterion described below, the final analytic sample comprised 169 participants.

This study was approved by the Institutional Review Board of the University of Utah (IRB #00149365). All participants provided written informed consent, obtained electronically, prior to enrollment. This study is reported in accordance with the STROBE statement^38^ and the TRIPOD+AI guideline.^28^ The protocol of the parent study has been published;^31^ no separate protocol was prepared for this analysis, and it was not registered.

### Outcome definition and inclusion criteria

Psychological distress was assessed daily using the BSI-18,^5^ administered via an end-of-day self-report survey. The BSI-18 comprises three six-item subscales—Somatization, Depression, and Anxiety—with each item rated on a 5-point Likert scale (0 = not at all to 4 = extremely). When multiple surveys were completed on the same day, only the last was retained, yielding one BSI-18 administration per participant-day.

Following BSI-18 scoring convention,^6,39^ a subscale with at most one missing item was prorated (mean of the available items scaled to six items); otherwise it was invalid. The Global Severity Index (GSI), the sum of the three prorated subscale scores, required all three to be valid. Only records with a valid GSI were retained, yielding 1,062 participant-day records from 169 participants.

GSI scores were then converted to sex-specific, normalized T-scores (mean = 50, standard deviation = 10) using published general-population norms.^40^ Psychological caseness was defined as a T-score ≥63, following the threshold recommended by Derogatis.^5^ The overall prevalence across the 1,062 participant-day records was 11.2% (male: 12.9%; female: 10.9%).

### Predictive task and feature extraction

The modeling objective was to predict daily psychological distress, operationalized as BSI-18 GSI caseness (T-score ≥63), for each participant-day record. Features were extracted from two sources: (1) baseline survey and demographic data collected prior to the monitoring period, and (2) passive sensor data with timestamps strictly preceding the end-of-day survey on each calendar day.

For each participant-day record, the considered window of sensor data was defined as the interval ending at the time of end-of-day survey completion and beginning at either (a) 24 hours prior to the survey, or (b) the time of the previous day’s end-of-day survey, whichever was more recent. This prevented double-counting across consecutive records.

The considered window was partitioned into 96 equally spaced 15-minute bins. Within each bin, raw sensor observations were aggregated directly at four temporal resolutions—15-minute, 1-hour, 6-hour, and 24-hour—and broadcast onto the shared 96-bin grid. This multi-resolution strategy was adopted because the temporal scale relevant to daily psychological distress is not known *a priori*. Accelerometer features included step count, walking duration, cadence, and acceleration magnitude statistics. GPS features included displacement, path length, and speed.

Of the 169 participants, 156 (92.3%) reported their home address. For these participants, distance-from-home and at-home indicator features were derived from GPS coordinates relative to the reported address. For the remaining participants, these features were treated as missing. Power state features captured screen lock/unlock counts, screen-on and plugged fractions, and battery level. Rate-of-change features were computed as the difference from the nearest prior observed bin. Per-bin data availability indicators were included as features to account for differential sensor coverage across participants and time.

Further, ten daily summary features were derived from accelerometer data over the full considered window, capturing sleep metrics (e.g., sleep period count, longest sleep duration) and activity summaries (e.g., active minutes per observed hour, step rate, peak cadence). Clock time was encoded via sinusoidal transformation, and day of week was encoded as a binary weekend indicator. Baseline survey variables were included after one-hot encoding of nominal categorical variables. The daily summary, weekend, and survey features, henceforth collectively termed *static* features, were constant within a given participant-day record. All 564 features are defined in Supplementary Table 1.

### Data preprocessing

The 1,062 participant-day records were first partitioned into non-overlapping sets using stratified group k-fold splitting, which preserved approximate class prevalence across sets while constraining all records from a given participant to fall exclusively into one set.

A 5-fold outer split was first applied. One fold was designated as the held-out test set (*n* = 210) and was excluded from all model development. The remaining 852 records underwent a separate 4-fold stratified group split for hyperparameter optimization, with three folds used for training and the held-out fold for validation in each iteration.

For final model retraining, the 852 non-test records were re-split into a training set (*n* = 730) and a validation set (*n* = 122) at a ratio of approximately 6:1. This training ratio maximized training data while retaining sufficient validation records for checkpoint selection and calibration.

The preprocessing pipeline comprised three steps, each fitted on the training partition and applied identically to the validation and test partitions to prevent data leakage. First, features that were constant or entirely missing in the training partition were removed from the initial set of 564 features. Second, continuous features were winsorized at ±3 standard deviations from the training mean to limit outlier influence. Third, all features were scaled to the unit interval [0, 1] via min–max normalization to ensure numerical stability.

The preprocessed data were then formatted differently for two model families. For classical machine learning (ML) algorithms, a two-dimensional representation was constructed by mean-aggregating each feature across 96 bins. Median imputation was applied to remaining missing values, with a binary missingness indicator appended for each imputed feature. For transformer models, the data were retained as a three-dimensional array (records × 96 bins × features) with missing values preserved for internal handling by the model’s masking mechanism.

### Modeling methods

Eight model configurations were evaluated. Four classical ML algorithms operated on the two-dimensional (mean-aggregated) representation, and two transformer architectures—a single-stream and a two-stream fusion variant—were each evaluated under two feature sets: unimodal (sensor features only) and multimodal (sensor and survey features combined), yielding four transformer configurations that operated on the three-dimensional temporal representation. All models used weighted binary cross-entropy loss with a cost-sensitive class weight multiplier included as a tunable hyperparameter to address class imbalance.

#### Classical machine learning models

Elastic net logistic regression (EN), linear support vector machine (SVM), extreme gradient boosting (XGB), and artificial neural network (ANN) were applied to the multimodal two-dimensional representation.

#### Single-stream standard transformer

A transformer classifier was developed for multivariate time-series data with missing values, processing each participant-day record as a sequence of 96 regularly spaced 15-minute bins. At each bin, observed feature values were projected into a dense embedding via a masked linear layer that gates missing values to zero. Two missingness handling strategies were explored as a hyperparameter (Supplementary Table 2). Temporal position was encoded through Time2Vec.^41^ The combined embedding was passed through transformer encoder layers^22^ with multi-head self-attention, followed by learned attention pooling to produce a single fixed-length representation per record, which was mapped to a binary logit via a linear classification head. Training used the AdamW optimizer.^42^

#### Two-stream fusion transformer

The attention-pooling layer can dilute the contribution of time-invariant features. Thus, the fusion architecture routes static and temporal features through separate encoding streams. Temporal features follow the same pipeline as in the single-stream model. Static features are extracted at a single bin position and encoded via an additive mask-embedding projection followed by a multilayer perceptron. The two representations are concatenated and passed to a linear classification head for intermediate fusion, trained end-to-end. The static stream contained only daily activity summaries and the weekend indicator under the unimodal configuration, and additionally included baseline survey features under the multimodal configuration.

### Hyperparameter optimization and model retraining

For each model configuration, hyperparameters were optimized via grid search over the 4-fold internal cross-validation described above. The configuration with the highest mean area under the receiver operating characteristic curve (AUROC) across the four folds was selected, with ties broken by lower expected calibration error (ECE). The full hyperparameter grids are provided in Supplementary Table 2.

The selected configuration was retrained on the full training set (*n* = 730) with four random seeds (0, 1, 2, 42), with the validation set (*n* = 122) used to select the transformer checkpoint with the lowest validation loss over 300 epochs.

Predicted probabilities on the held-out test set were calibrated using Platt scaling,^43^ which was chosen as the primary calibration method due to its stability on small validation sets. Isotonic regression and temperature scaling^44^ were evaluated as secondary methods and reported in Supplementary Table 3.

### Performance measurement

All metrics were computed on the held-out test set for a seed ensemble of each model: the Platt-scaled probabilities of the four seed models were averaged for each record, and each metric was computed once. Confidence intervals (95% CIs) were obtained from 2,000 bootstrap samples of the 34 test participants, resampled with replacement together with all their records. Discrimination was assessed with the AUROC and the area under the precision–recall curve (AUPRC). Calibration was assessed with the Brier score,^45^ log loss, calibration intercept and slope, and ECE.^44^ Classification was assessed with sensitivity, specificity, positive predictive value (PPV; precision), negative predictive value (NPV), F1 score, balanced accuracy, Matthews correlation coefficient (MCC) and Youden index, at thresholds of 0.50 and 0.12, approximating the test-set outcome prevalence (11.9%). Clinical utility was assessed with decision-curve analysis,^27,46^ using net benefit standardized by the outcome prevalence. Complete metric definitions and additional results are reported in Supplementary Table 3.

### Feature importance

Feature importance was assessed using SHAP (SHapley Additive exPlanations),^30^ implemented via the GradientSHAP method in the Captum library.^47^ Attributions were computed for the multimodal fusion transformer—the primary model of interest—independently for each of the four retrained seed models on the held-out test set and averaged elementwise across seeds to produce a stable importance estimate.

### Intraclass correlation analysis

Intraclass correlation coefficients (ICCs), the proportion of total variance attributable to between-participant differences, were computed for the binary outcome and passive sensor features. For the binary distress outcome, a logistic generalized linear mixed model with a random intercept for participant was fitted and the ICC derived on the latent scale. For continuous sensor features, linear random-intercept models were fitted using restricted maximum likelihood estimation. Epoch-level features were aggregated to record level by averaging across epochs, preserving records with partial missingness.

## Results

### Participants and outcome

The 169 participants (Table 1) contributed a median of 7 records each (interquartile range 6–7). They were mostly young (median age 27 years), female (86.4%) and White (82.2%), and 65 (38.5%) were full-time students.

**Table 1.** Demographic characteristics of study participants. Demographic characteristics of the 169 unique participants included in the study. Age is summarized using descriptive statistics, and categorical variables are presented as counts and percentages. Participants were allowed to select multiple racial identities; therefore, the sum of race category percentages may exceed 100%.

| **Age (years)** |  |
|---|---|
| *Mean* | 31.70 |
| *Standard deviation* | 13.12 |
| *Minimum* | 18.00 |
| *Maximum* | 76.00 |
| *Median* | 27.00 |
| **Biological sex** |  |
| *Female* | 146 (86.39%) |
| *Male* | 23 (13.61%) |
| **Gender** |  |
| *Women* | 141 (83.43%) |
| *Men* | 21 (12.43%) |
| *Others / did not respond* | 7 (4.14%) |
| **Race** |  |
| *White* | 139 (82.25%) |
| *Asian* | 22 (13.02%) |
| *Black or African American* | 7 (4.14%) |
| *American Indian or Alaska Native* | 6 (3.55%) |
| *Native Hawaiian or Other Pacific Islander* | 2 (1.18%) |
| Others | 4 (2.37%) |
| Did not respond | 4 (2.37%) |
| **Ethnicity** |  |
| *Not Hispanic or Latino* | 140 (82.84%) |
| *Hispanic or Latino* | 27 (15.98%) |
| Did not respond | 2 (1.18%) |

The held-out test set contained 210 participant-day records (25 cases; 11.9%) from 34participants, and the development data 852 records (94 cases; 11.0%) from 135 participants.

### Discrimination

Table 2 and Figure 1 summarize test-set performance after Platt scaling. The multimodal fusion transformer (AUROC 0.84; 95% CI 0.69–0.95) and the SVM (0.84; 0.71–0.93) had the highest AUROC, followed by XGB (0.82; 0.63–0.94) and the remaining three multimodal models (0.79– 0.81). The confidence intervals of these six multimodal models (the four classical models and the two multimodal transformers) overlapped substantially. The two unimodal transformers did not discriminate better than chance (AUROC 0.50 and 0.49; 95% CIs 0.33–0.74 and 0.26–0.77).

**Figure 1.**
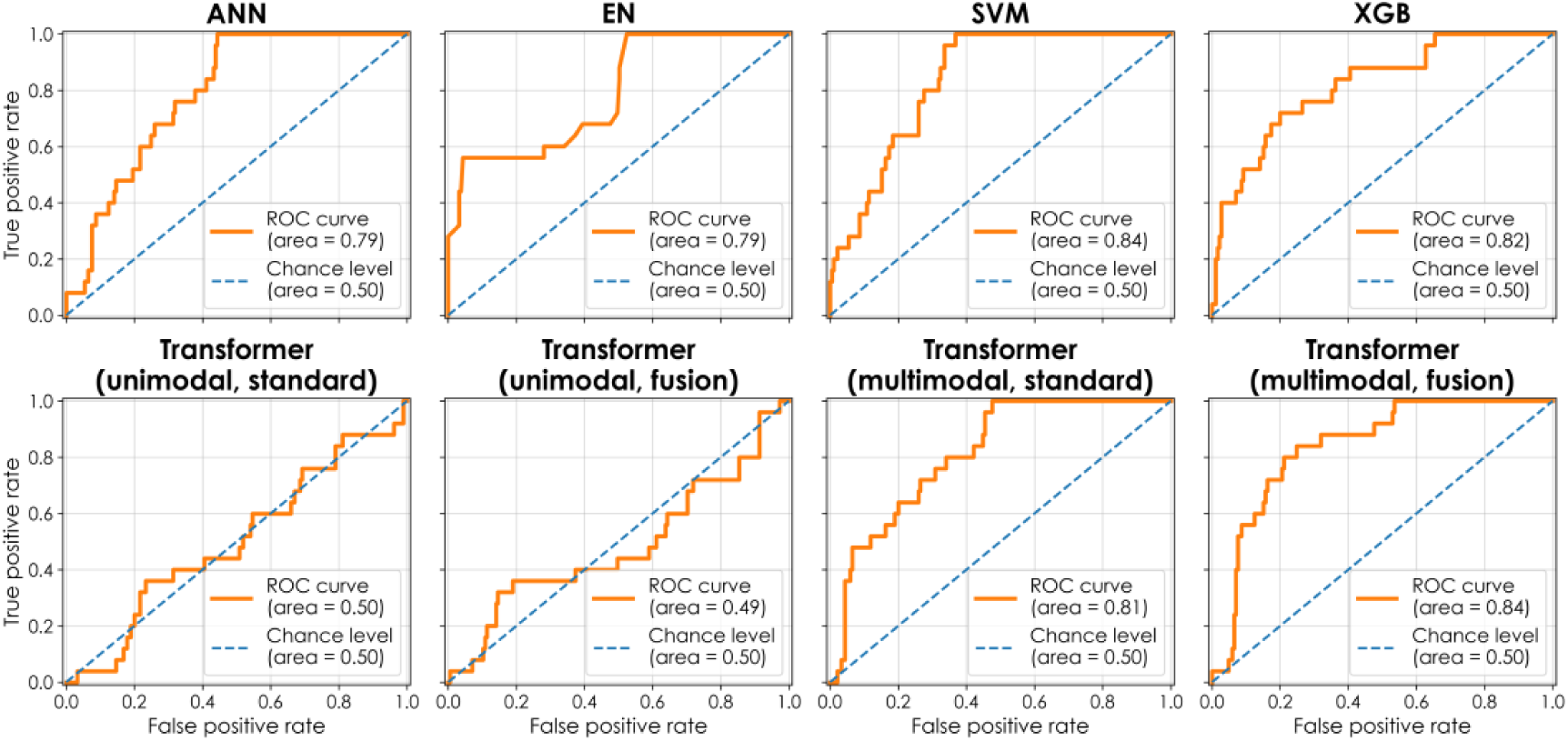
Area under the receiver operating characteristic curve. The area under the receiver operating characteristic curve (ROC, orange solid line) for the eight prediction models: artificial neural network (ANN), elastic net (EN), support vector machine (SVM), extreme gradient boosting (XGB), and four transformer models with unimodal or multimodal data input and standard or fusion architecture. All metrics were evaluated on the held-out test set (*n* = 210) with probability estimates averaged across four random-seeded models (0, 1, 2, 42) used during model retraining after hyperparameter optimization. Predicted probability estimates of each model have been calibrated by Platt scaling.

**Table 2.** Performance of models predicting daily psychological distress. Performance metrics (best bolded in red) are shown for the eight prediction models: artificial neural network (ANN), elastic net (EN), support vector machine (SVM), extreme gradient boosting (XGB), and four transformer models with unimodal or multimodal data input and standard or fusion architecture. All metrics were evaluated on the held-out test set (n = 210) with probability estimates averaged across four random-seeded models (0, 1, 2, 42) used during model retraining after hyperparameter optimization. Predicted probability estimates of each random-seeded model have been calibrated by Platt scaling. The 95% confidence intervals (in parentheses) were computed by cluster bootstrapping (2,000 rounds) of the 34 unique participants in the test set. Cells shown as N/A represent undefined division-by-zero values. Threshold-dependent statistics were evaluated at the decision thresholds of 0.50 and 0.12; the latter approximates the outcome prevalence in the held-out test set (11.9%). See Supplementary Table 3 for a fuller list of performance statistics and their confidence intervals computed via cluster bootstrapping. AUPRC = area under the precision–recall curve; AUROC = area under the receiver operating characteristic curve; Cal. = calibration; ECE = expected calibration error; Bal. accuracy = balanced accuracy; MCC = Matthews correlation coefficient; NPV = negative predictive value.

| **Algorithm** | **ANN** | **EN** | **SVM** | **XGB** | **Unimodal transformer** |  | **Multimodal transformer** |  |
|---|---|---|---|---|---|---|---|---|
|  |  |  |  |  | **Standard** | **Fusion** | **Standard** | **Fusion** |
| *Threshold-independent statistics* |  |  |  |  |  |  |  |  |
| AUPRC | 0.32<br>(0.15, 0.51) | **0.54**<br>(0.12, 0.86) | 0.43<br>(0.13, 0.69) | 0.44<br>(0.09, 0.75) | 0.12<br>(0.05, 0.23) | 0.13<br>(0.06, 0.30) | 0.31<br>(0.10, 0.68) | 0.35<br>(0.17, 0.74) |
| AUROC | 0.79<br>(0.67, 0.90) | 0.79<br>(0.56, 0.94) | 0.84<br>(0.71, 0.93) | 0.82<br>(0.63, 0.94) | 0.50<br>(0.33, 0.74) | 0.49<br>(0.26, 0.77) | 0.81<br>(0.64, 0.94) | **0.84**<br>(0.69, 0.95) |
| Brier score | 0.10<br>(0.05, 0.15) | 0.10<br>(0.06, 0.15) | 0.11<br>(0.06, 0.16) | **0.09**<br>(0.05, 0.13) | 0.11<br>(0.05, 0.18) | 0.11<br>(0.05, 0.18) | 0.11<br>(0.06, 0.17) | 0.09<br>(0.05, 0.14) |
| Cal. intercept | -0.16<br>(-1.24, 0.61) | -0.88<br>(-1.79, -0.11) | -0.93<br>(-1.91, -0.15) | -0.48<br>(-1.47, 0.24) | 0.16<br>(-0.89, 0.86) | **0.08**<br>(-0.96, 0.77) | -0.78<br>(-2.00, 0.14) | -0.36<br>(-1.40, 0.43) |
| Cal. slope | 0.86<br>(0.54, 1.51) | 0.93<br>(0.23, 2.55) | 0.83<br>(0.48, 1.45) | 0.83<br>(0.30, 1.40) | 0.06<br>(-1.63, 3.20) | 0.08<br>(-5.02, 5.73) | 0.68<br>(0.18, 1.39) | **1.00**<br>(0.48, 2.89) |
| ECE | **0.05**<br>(0.04, 0.14) | 0.15<br>(0.09, 0.21) | 0.10<br>(0.06, 0.18) | 0.07<br>(0.04, 0.13) | 0.07<br>(0.05, 0.16) | 0.07<br>(0.06, 0.17) | 0.10<br>(0.06, 0.18) | 0.05<br>(0.05, 0.15) |
| Log loss | 65.18<br>(38.67, 97.04) | 68.85<br>(47.00, 95.17) | 66.20<br>(42.08, 93.30) | 62.76<br>(41.30, 88.61) | 78.06<br>(43.91, 122.76) | 77.02<br>(44.86, 118.01) | 72.09<br>(44.33, 106.57) | **62.46**<br>(38.71, 89.63) |
| *Threshold-dependent statistics (evaluated at threshold = 0.50)* |  |  |  |  |  |  |  |  |
| Bal. accuracy | 0.51<br>(0.48, 0.56) | **0.76**<br>(0.52, 0.91) | 0.64<br>(0.51, 0.78) | 0.68<br>(0.49, 0.83) | 0.50<br>(0.50, 0.50) | 0.50<br>(0.50, 0.50) | 0.69<br>(0.49, 0.87) | 0.50<br>(0.48, 0.55) |
| F1 | 0.11<br>(0.00, 0.19) | **0.60**<br>(0.11, 0.84) | 0.36<br>(0.12, 0.54) | 0.49<br>(0.00, 0.74) | 0.00<br>(0.00, 0.00) | 0.00<br>(0.00, 0.00) | 0.44<br>(0.06, 0.74) | 0.06<br>(0.00, 0.15) |
| MCC | 0.04<br>(-0.06, 0.17) | **0.55**<br>(0.14, 0.82) | 0.26<br>(0.02, 0.47) | 0.45<br>(-0.03, 0.70) | N/A | N/A | 0.36<br>(-0.02, 0.70) | -0.01<br>(-0.07, 0.09) |
| NPV | 0.88<br>(0.80, 0.96) | **0.94**<br>(0.88, 0.99) | 0.92<br>(0.84, 0.98) | 0.92<br>(0.86, 0.97) | 0.88<br>(0.79, 0.95) | 0.88<br>(0.79, 0.95) | 0.93<br>(0.86, 0.98) | 0.88<br>(0.79, 0.96) |
| Precision | 0.17<br>(0.00, 0.58) | **0.64**<br>(0.14, 1.00) | 0.32<br>(0.09, 0.59) | 0.63<br>(0.00, 0.89) | N/A | N/A | 0.41<br>(0.07, 0.83) | 0.11<br>(0.00, 0.14) |
| Recall | 0.08<br>(0.00, 0.20) | **0.56**<br>(0.09, 0.87) | 0.40<br>(0.13, 0.67) | 0.40<br>(0.00, 0.70) | 0.00<br>(0.00, 0.00) | 0.00<br>(0.00, 0.00) | 0.48<br>(0.06, 0.84) | 0.04<br>(0.00, 0.17) |
| Specificity | 0.95<br>(0.87, 1.00) | 0.96<br>(0.88, 1.00) | 0.89<br>(0.79, 0.96) | 0.97<br>(0.94, 0.99) | **1.00**<br>(1.00, 1.00) | 1.00<br>(1.00, 1.00) | 0.91<br>(0.81, 0.99) | 0.96<br>(0.88, 1.00) |
| Youden index | 0.03<br>(-0.04, 0.11) | **0.52**<br>(0.05, 0.83) | 0.29<br>(0.03, 0.55) | 0.37<br>(-0.02, 0.66) | 0.00<br>(0.00, 0.00) | 0.00<br>(0.00, 0.00) | 0.39<br>(-0.02, 0.74) | 0.00<br>(-0.04, 0.09) |
| *Threshold-dependent statistics (evaluated at threshold = 0.12)* |  |  |  |  |  |  |  |  |
| Bal. accuracy | 0.70<br>(0.58, 0.84) | 0.61<br>(0.42, 0.79) | **0.81**<br>(0.73, 0.88) | 0.73<br>(0.55, 0.85) | 0.54<br>(0.39, 0.73) | 0.52<br>(0.39, 0.73) | 0.73<br>(0.55, 0.88) | 0.74<br>(0.58, 0.85) |
| F1 | 0.35<br>(0.17, 0.52) | 0.27<br>(0.09, 0.46) | **0.41**<br>(0.21, 0.59) | 0.38<br>(0.13, 0.59) | 0.21<br>(0.06, 0.36) | 0.19<br>(0.04, 0.35) | 0.38<br>(0.16, 0.59) | 0.36<br>(0.15, 0.56) |
| MCC | 0.27<br>(0.09, 0.44) | 0.14<br>(-0.11, 0.37) | **0.40**<br>(0.26, 0.54) | 0.31<br>(0.05, 0.52) | 0.05<br>(-0.15, 0.27) | 0.03<br>(-0.17, 0.27) | 0.31<br>(0.06, 0.53) | 0.31<br>(0.09, 0.49) |
| NPV | 0.95<br>(0.88, 1.00) | 0.93<br>(0.82, 1.00) | **1.00**<br>(1.00, 1.00) | 0.96<br>(0.90, 0.99) | 0.89<br>(0.78, 0.98) | 0.89<br>(0.77, 0.98) | 0.96<br>(0.87, 1.00) | 0.97<br>(0.91, 1.00) |
| Precision | 0.22<br>(0.10, 0.38) | 0.16<br>(0.05, 0.32) | **0.26**<br>(0.11, 0.42) | 0.25<br>(0.08, 0.46) | 0.14<br>(0.04, 0.27) | 0.13<br>(0.03, 0.26) | 0.25<br>(0.10, 0.45) | 0.22<br>(0.09, 0.40) |
| Recall | 0.76<br>(0.56, 1.00) | 0.72<br>(0.33, 1.00) | **1.00**<br>(1.00, 1.00) | 0.76<br>(0.39, 0.96) | 0.40<br>(0.13, 0.77) | 0.36<br>(0.09, 0.75) | 0.76<br>(0.40, 1.00) | 0.88<br>(0.57, 1.00) |
| Specificity | 0.64<br>(0.49, 0.78) | 0.50<br>(0.33, 0.68) | 0.61<br>(0.46, 0.75) | **0.70**<br>(0.59, 0.80) | 0.67<br>(0.56, 0.78) | 0.68<br>(0.57, 0.80) | 0.69<br>(0.54, 0.83) | 0.59<br>(0.43, 0.75) |
| Youden index | 0.40<br>(0.15, 0.69) | 0.22<br>(-0.16, 0.58) | **0.61**<br>(0.46, 0.75) | 0.46<br>(0.09, 0.69) | 0.07<br>(-0.22, 0.45) | 0.04<br>(-0.22, 0.45) | 0.45<br>(0.09, 0.76) | 0.47<br>(0.16, 0.70) |
| *Other statistics* |  |  |  |  |  |  |  |  |
| Number of model parameters | 782,337 | 1,014 | 1,014 | 1,576 | 948,995 | 177,091 | 737,283 | 138,371 |
| Average per-model training time (in seconds) | 6.98 | 1.07 | 0.05 | 0.26 | 96.43 | 61.82 | 133.82 | 83.63 |

With a test-set prevalence of 11.9%, a non-informative model has an AUPRC of approximately 0.12. EN had the highest AUPRC (0.54; 95% CI 0.12–0.86), and the unimodal transformers were at the prevalence level (0.12 and 0.13). EN ranked about half of the cases above almost all non-cases but separated the rest poorly (Figure 1), which explains its high AUPRC but mid-ranking AUROC.

### Calibration and overall accuracy

After Platt scaling, XGB (0.09) and the multimodal fusion transformer (0.09) had the lowest Brier scores, against 0.10 for a prevalence-only model; the other models scored 0.10–0.11, close to this reference. Calibration slopes were 0.83–1.00 for the multimodal models except the multimodal standard transformer (0.68, overconfident), with wide CIs. The calibration intercepts (−0.16 to −0.93) of the multimodal models showed that they tended to overestimate risk; the CIs excluded the null value only for EN and SVM.

In sensitivity analyses (Supplementary Table 3), temperature scaling improved intercepts but gave underconfident slopes, and isotonic regression changed AUROC by −0.10 to +0.02, with the largest drop for the SVM (0.84 to 0.74).

### Classification at two decision thresholds

The choice of threshold changed both the absolute performance and the ranking of the models (Table 2). At 0.50, the models flagged 0–15% of test records, and the unimodal transformers predicted no cases. EN was the best classifier, with the highest sensitivity and PPV of any model: it identified 14 of 25 cases (sensitivity 0.56, specificity 0.96, PPV 0.64). XGB had a similar PPV (0.63) but a lower sensitivity (0.40), and the multimodal standard transformer had the next-highest sensitivity (0.48) but a lower PPV (0.41). The ANN and the multimodal fusion transformer detected only 2 and 1 cases, respectively. Only EN and XGB had a positive net benefit at this threshold (Figure 2).

**Figure 2.**
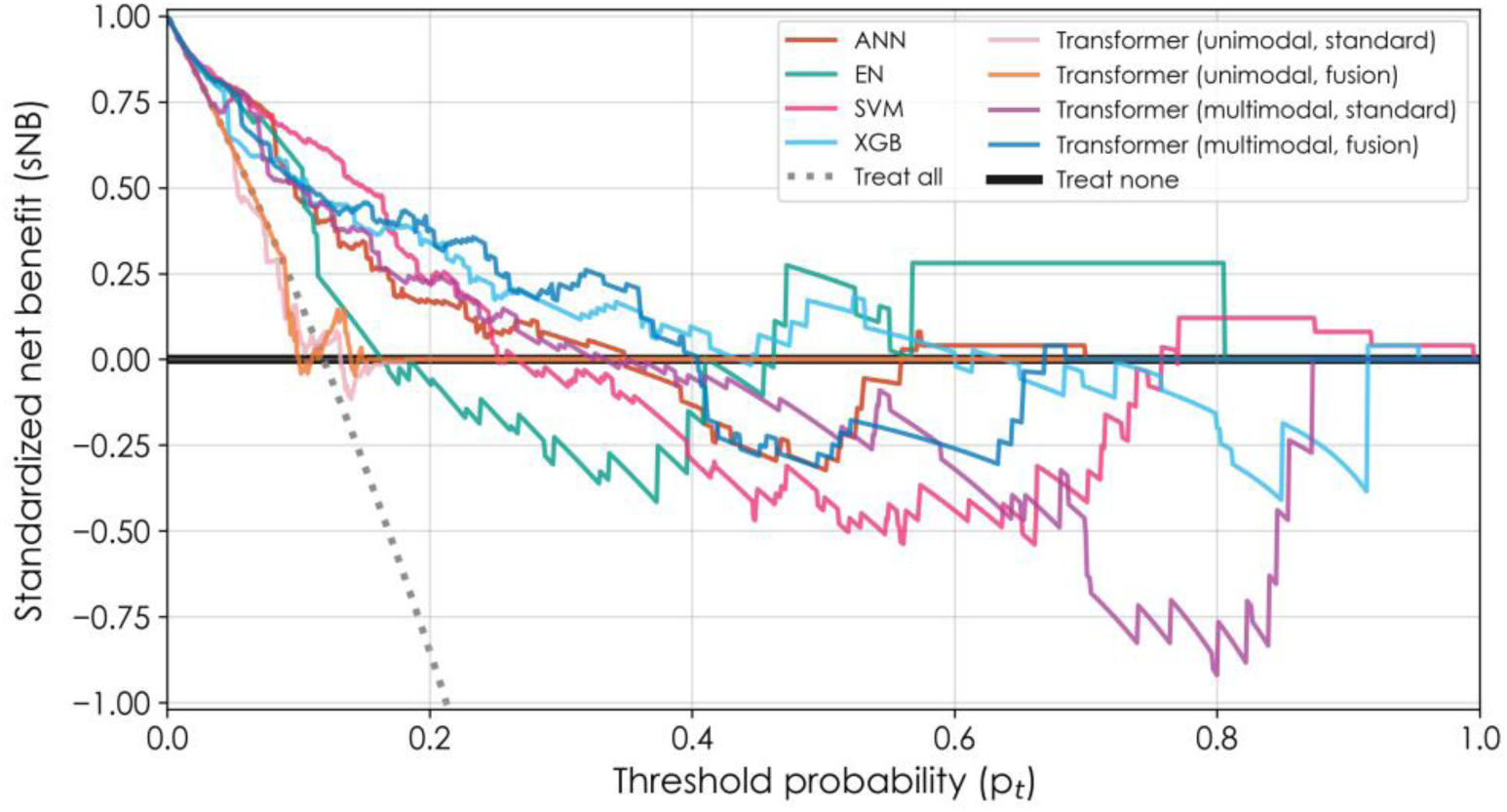
Decision-curve analysis of models predicting daily psychological distress. Standardized net benefit (sNB) is evaluated on the held-out test set (*n* = 210) with probability estimates averaged across four random-seeded models (0, 1, 2, 42) used during model retraining after hyperparameter optimization. It is shown as a function of the decision threshold probabilities (p*_t_*) for the eight prediction models: artificial neural network (ANN), elastic net (EN), support vector machine (SVM), extreme gradient boosting (XGB), and four transformer models with unimodal or multimodal data input and standard or fusion architecture. The treat-all strategy (gray dotted line) represents the sNB obtained by assigning the intervention to all records regardless of predicted risk, while the treat-none strategy (solid black horizontal line) corresponds to withholding intervention from all records (sNB = 0).

At 0.12, the models flagged 32–52% of records. The multimodal models had a sensitivity of 0.72–1.00, a specificity of 0.50–0.70, an NPV of 0.93–1.00 and a PPV of 0.16–0.26, that is, three to five false-positive records per detected case. The SVM identified all 25 cases (NPV 1.00) while flagging 46% of records. EN, the best model at 0.50, was the weakest multimodal model at 0.12, with the lowest sensitivity, specificity, NPV and PPV.

### Clinical utility

Figure 2 shows the decision curves. For threshold probabilities between 0.05 and 0.25, the SVM, XGB, ANN and both multimodal transformers had a higher standardized net benefit than both default strategies (treat-all; treat-none). At 0.12, the SVM had the highest standardized net benefit. Above about 0.40, most curves became unstable and often fell below treat-none, consistent with the few cases that received high predicted risks. The unimodal transformers gave no net benefit over treat-none above about 0.15.

### Model complexity

EN, SVM and XGB had 1,014–1,576 parameters and trained in ≤1.1 s; the ANN had 782,337 parameters and took 7.0 s, and the transformers had 138,371–948,995 parameters and took 62– 134 s (Table 2).

### Feature importance

In the multimodal fusion transformer, survey features had the largest SHAP values (absolute values up to about 0.15; Figure 3). Four of the five highest-ranked survey features measured functional impairment summarized from the Life Functioning Questionnaire^37^ (difficulty socializing with friends, socializing with family, fulfilling work or school duties, and completing household duties without conflict), and higher reported difficulty increased predicted risk. For the household-duties item, the SHAP values formed four clusters, one for each ordinal response level, and rose with each higher level of difficulty. Full-time student status decreased predicted risk. The most important daily and temporal sensor features had SHAP values mostly within ±0.04. More sustained-activity bouts and active minutes per observed hour were associated with higher predicted risk, and more steps per observed hour, phone use and a higher screen-on fraction with lower predicted risk. Two of the ten sensor features shown measured accelerometer data coverage (daily and 6-hour) rather than behavior.

**Figure 3.**
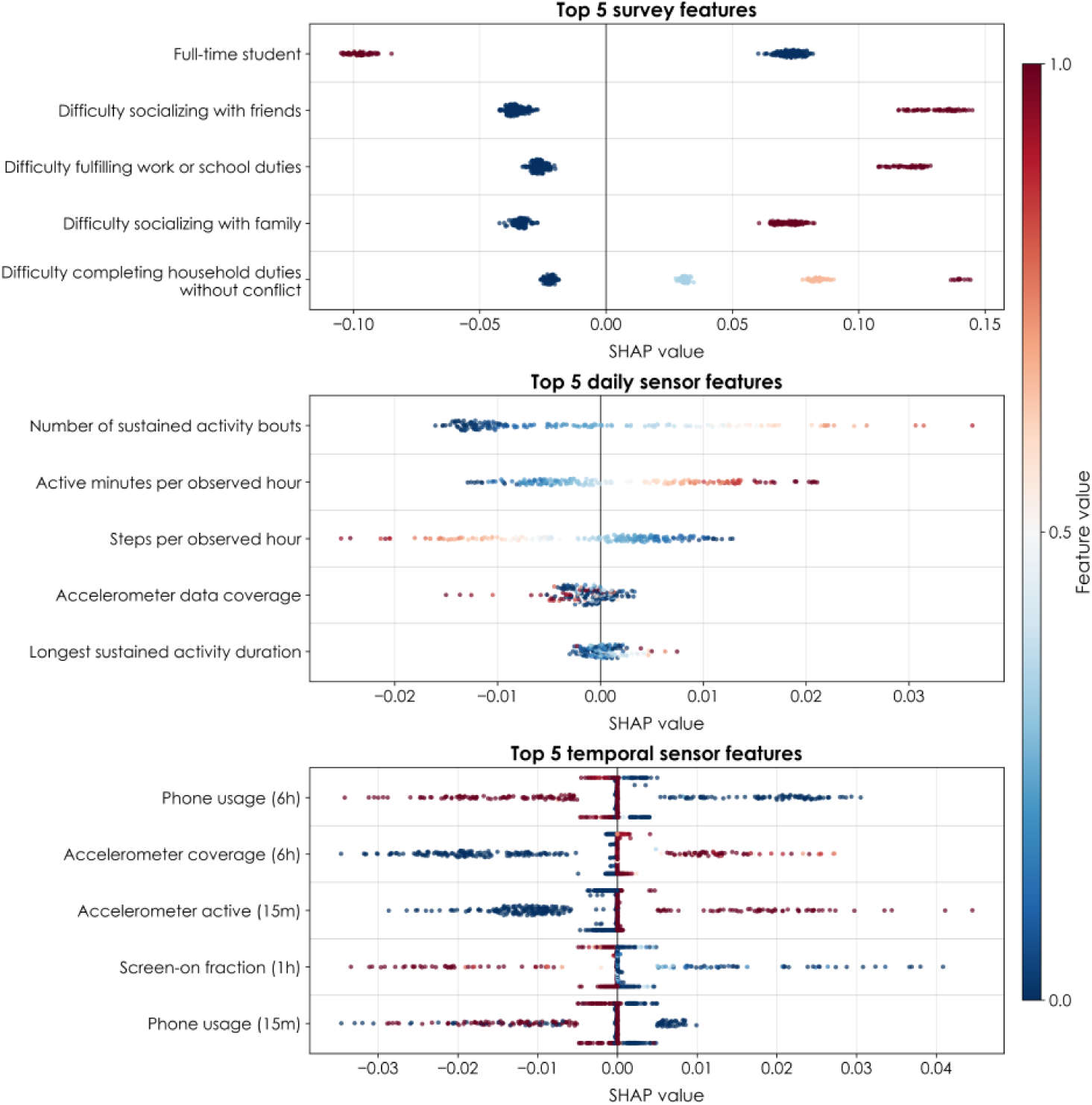
Feature importance for the top five survey, daily sensor, and temporal sensor features returned by the multimodal fusion transformer. The beeswarm plots show the SHAP values across the 96 15-minute time bins and all records of the top five survey (top panel), daily sensor (middle panel), and temporal sensor (bottom panel) features (by mean absolute SHAP values across test-set records and timestamps) according to the multimodal fusion transformer evaluated on the held-out test set (*n* = 210). Min–max normalized feature values were represented by the color of each data point (red for high values and blue for low values). The color distribution indicates whether the feature is a risk or protective factor. For example, being a full-time student (first survey feature) had high feature values (red) in the negative regime of SHAP values, indicating that it is a protective factor of daily psychological distress.

## Discussion

### Principal findings

Three findings emerged. First, compact classical models (EN, SVM, XGB) performed as well as the best transformer: the SVM matched its AUROC, EN had the highest AUPRC and XGB the lowest Brier score (Table 2). Second, the transformers that received sensor data alone performed at chance; with survey data added, their AUROC reached that of the classical models. Third, baseline self-report, chiefly functional-impairment items, dominated the predictions, and sensor features contributed small effects of mixed direction (Figure 3). The predictive signal in these data therefore comes mainly from self-report; the added value of sensing was not tested directly.

### Classical versus transformer models

The similar performance of compact and deep models agrees with a systematic review that found no benefit of machine learning over logistic regression in clinical prediction comparisons at low risk of bias^25^ and with evidence that tree ensembles outperform deep networks on medium-sized tabular datasets.^26^ With about 1,000 records and 10⁵–10⁶ parameters, the transformers were heavily overparameterized. The *fusion* architecture handled this better: it had about one-fifth as many parameters as the standard multimodal transformer and better point estimates for every threshold-independent metric, including calibration slope (1.00 vs 0.68). Even so, it performed no better than the SVM or XGB, with overlapping CIs and 300–1,700× more training time (Table 2). The classical models received each feature as a daily mean without within-day timing; because they still matched the transformers, within-day temporal patterns appear to add little in these data. Providing the classical models with each feature per 15-minute bin would have multiplied the feature count 96-fold relative to about 1,000 records. The transformers could offer more detailed explanations. SHAP can explain individual records for any model, but only the transformers can show when during the day a feature contributed: for the fusion transformer, SHAP values were computed for each 15-minute bin of each record (pooled across bins in Figure 3), whereas the classical models, which received daily means, can be explained only at the whole-day level. At the current data scale, the simpler models are preferable for deployment and transparency.

### Contribution of passive sensing

The chance-level performance of the sensor-only transformers is an important negative result. It is consistent with reviews reporting small samples, heterogeneous features and limited reproducibility^11,15^ and with poor generalization across populations and datasets.^16,17^ Several explanations apply: (i) between-person differences in phone and activity habits can hide within-person changes that indicate distress; (ii) the less-than-24-hour sensor window per record may be too short to capture slow behavioral changes; (iii) only 94 development cases are few for a transformer learning temporal patterns across 96 time bins; and (iv) all sensor data came from the phone, which is not always carried, so activity measures may partly reflect phone-carrying habits. The two data-coverage features among the top ten (Figure 3) may reflect engagement with the phone and the study protocol,^20^ which itself relates to symptom scores,^19^ rather than distress. For steps per observed hour, high values were associated with lower predicted risk, whereas medium to zero values were indeterminate: they could reflect either low physical activity or a phone that was not being carried. We therefore consider the sensor-level SHAP patterns hypothesis-generating only; SHAP values describe the behavior of the model,^30^ not causal effects.

### Interpretation of survey predictors

Four of the five most important survey predictors were baseline items on difficulty in social, occupational and household functioning (Figure 3). These items were collected a median of 28 days before the sensing week. The model therefore does not simply re-measure the outcome at the same time point. However, functional impairment is closely related to daily psychological distress, so a strong association between baseline impairment and later distress is expected.

Distress was strongly clustered within participants. The 119 case-days came from 49 of the 169 participants (29%), and in a random-intercept logistic model the intraclass correlation coefficient (ICC) of caseness was 0.64: about two-thirds of the variation in the propensity for distress lay between participants and one-third within participants from day to day. Since most of the variance was driven by between-person differences, this observation supports splitting and bootstrapping the data at the participant level. Most sensor features varied more from day to day than between participants (median ICC of the daily feature values 0.42; range 0.02–1.00): GPS movement features, such as maximum speed, varied mostly within participants (ICC 0.02–0.12), whereas accelerometer sample counts, a measure of data coverage, were largely stable within participants (ICC 0.90–0.92).

Because the survey features are fixed for each participant, they can only separate *participants* with a high or low tendency to distress. They cannot tell which of a person’s 6–7 days are distressed days. Only the sensor features vary from day to day, and those features had little predictive value on their own. Most of the discrimination in the multimodal models is therefore probably *between-person*: the models identify who is at risk, not when. This is useful for screening, because a few impairment items are much shorter than repeated BSI-18 assessments. However, it limits the claim that the models detect distress *passively* or in real time.

### Choice of threshold and calibration

The two thresholds correspond to two clinical uses, and the best model depended on the use. At 0.50, the multimodal models worked as *high-specificity* classifiers but missed 44–96% of cases. This threshold treats a false positive as equally harmful as a missed case, which may suit an expensive or burdensome intervention but not screening; accordingly, net benefit at this threshold was small or negative.

At 0.12, the multimodal models worked as *high-sensitivity* classifiers with high NPV but low PPV. They are therefore best suited to *ruling out* distress or to *triaging* people for a low-cost confirmatory step, such as the full BSI-18 or a clinician check-in, at the cost of flagging more than a third of records. Decision-curve analysis supports this use: the better models exceeded both default strategies at thresholds of 0.05–0.25, where a false positive costs much less than a missed case.^27,46^

Because model rankings changed with the threshold, comparisons at a single threshold can mislead: EN’s concentration of high scores in a subgroup of clear cases was an advantage at 0.50 but a disadvantage at 0.12. Threshold selection is a clinical decision; until the intended setting is known, threshold-independent metrics and decision curves are more informative than a single confusion matrix.

Calibration is necessary for risk communication and threshold selection.^29^ Platt scaling tended to overestimate risk, possibly because it was fitted on only 122 validation records. Recalibration to the local prevalence before deployment is therefore necessary.

### Strengths and limitations

Strengths include the multimodal design and the comparison of eight model configurations under a common pipeline, with discrimination, calibration, classification and decision-curve analysis reported together with participant-level bootstrap CIs, as recommended.^28,45^

This study has several limitations. First, the test set contained only 25 cases among 210 participant-day records from 34 participants. The 95% CIs for the AUROC of the multimodal models were therefore 0.22–0.38 wide. We did not test differences between models formally, so we cannot conclude that any multimodal model is better than another. Second, the evaluation used one internal split with no external or temporal validation. Third, each participant contributed several records. The participant-level split prevents identity-based leakage,^21^ and the participant-level bootstrap accounts for the correlation between records of the same participant, but with so few test participants the CIs are themselves imprecise. Fourth, the sample was young, mostly female and White, and included many full-time students; it was recruited in one US state with risk-stratified sampling and excluded people with recent suicidal ideation, prior suicide attempts, mania or psychosis; and only iPhone users with sensor data were analyzed. These factors limit generalizability, particularly of PPV, NPV and calibration, because the case prevalence reflects the study design rather than the population. Fifth, caseness was based only on the GSI T-score criterion, a commonly used convention,^48^ and not on the full caseness rule (T-score ≥63 on the GSI or on at least two subscales),^5,40^ and the norms were derived from a 7-day, not a single-day, recall period. Sixth, with 564 candidate features and a small number of events, the sample size is below current recommendations for prediction-model development,^49^ which increases the risk of overfitting.

### Implications and future work

For clinical and public-health use, our results suggest that a simple, locally recalibrated classical model could serve as a first-line triage aid, and that a few functional-impairment items may carry most of its signal; passive sensing should be added only when it is shown to improve performance. Future work should quantify the incremental value of sensing over self-report, validate the models externally and prospectively in more diverse populations, and investigate within-person models and longer sensor windows.

## Conclusions

Compact classical machine-learning models (elastic net, SVM and XGB) identified clinically significant daily psychological distress as well as transformer models with about 90–730 times more parameters. Most of the predictive signal came from baseline survey items, mainly on functional impairment, and passive sensing alone did not discriminate cases. At a prevalence-based threshold, the best models gave high sensitivity and NPV with low PPV, which makes them candidates for low-burden rule-out triage. External validation and a formal test of the added value of sensing are necessary before clinical use.

## Data Availability

The data supporting the findings of this study are currently under review for deposition in a public repository. Until they become publicly available, the dataset may be obtained from Brian Baucom upon reasonable request. Code for development and reproducibility is publicly available at https://github.com/delacylab/DHI_BSI_Modeling.

## Acknowledgements

We thank the study participants. Computations were performed using resources at the Center for High Performance Computation at the University of Utah, which is partially funded by the NIH Shared Instrumentation Grant 1S10OD021644-01A1.

## Author contributions

IGS: Conceptualization, Formal analysis, Writing – original draft. WL: Methodology, Software, Formal analysis, Visualization, Writing – review and editing. REK: Data curation, Investigation, Writing – review and editing. BRWB: Funding acquisition, Investigation, Writing – review and editing. NdL: Conceptualization, Supervision, Writing – review and editing.

## Statements and Declarations

### Ethical considerations

This study was approved by the Institutional Review Board of the University of Utah, Salt Lake City, UT, USA (IRB #00149365; approval date March 28, 2022).

### Consent to participate

All participants provided written informed consent, obtained electronically, prior to enrollment.

### Consent for publication

Not applicable.

### Declaration of conflicting interest

The authors declare no potential conflicts of interest with respect to the research, authorship, and/or publication of this article.

### Funding statement

The author(s) received no financial support for the research, authorship, and/or publication of this article.

## Supplementary material

**Supplementary Figure 1.**
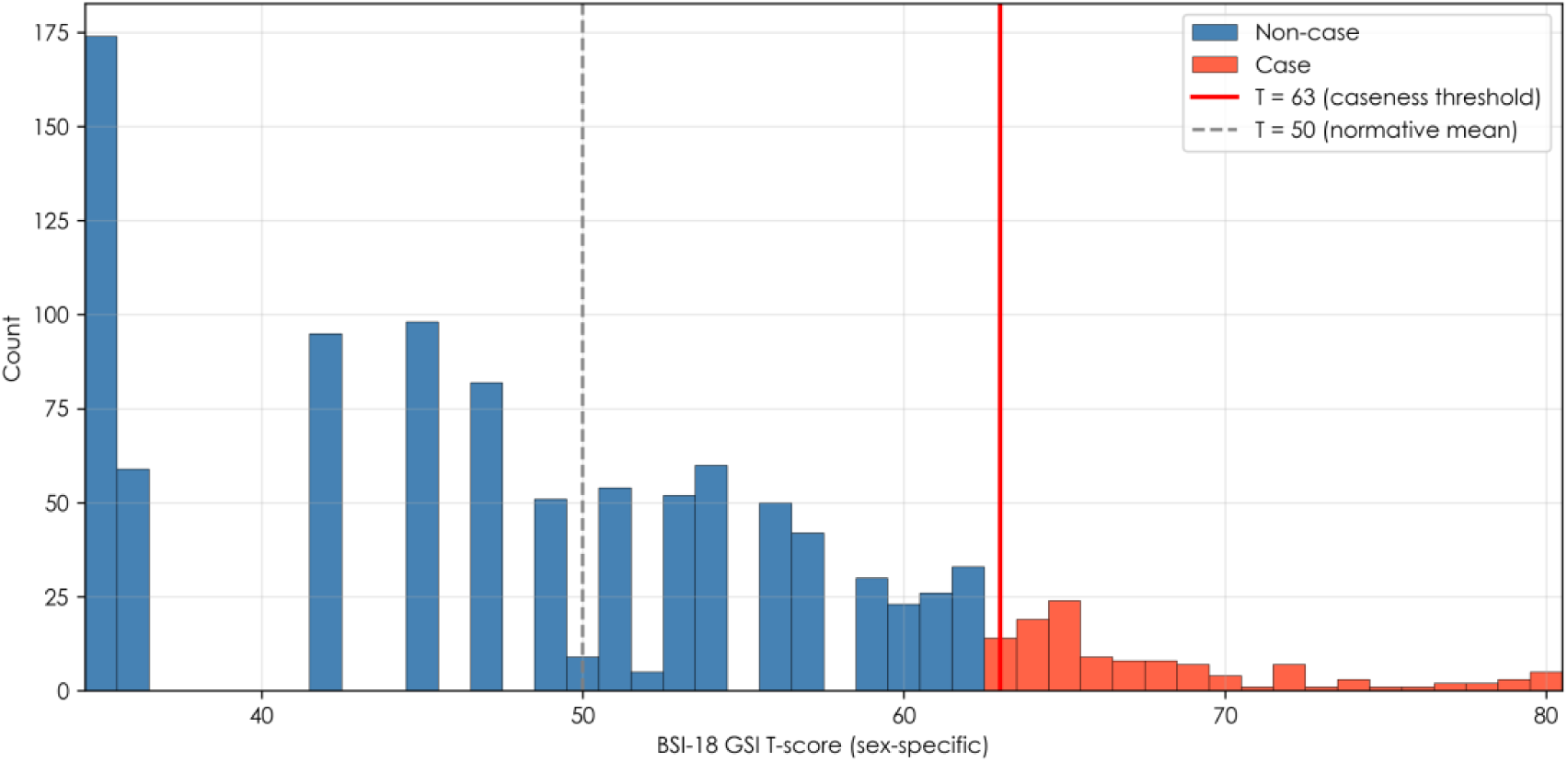
Distribution of BSI-18 Global Severity Index T-scores (sex-specific norms) across all records. Red bars: cases (T ≥63, red line); blue bars: non-cases. Dashed line: normative mean (T = 50).

## Notes

### Competing Interest Statement

The authors have declared no competing interest.

### Author Declarations

IRB #00149365 of University of Utah gave ethical approval for this work.

